# Enhancing Parental Knowledge of Child Safety: An Interventional Educational Campaign

**DOI:** 10.1101/2021.06.20.21259168

**Authors:** Mohamad-Hani Temsah, Fadi Aljamaan, Ali Alhaboob, Badr Almosned, Raghad Alsebail, Reem Temsah, Abdulrahman Senjab, Abdulrahman Alarfaj, Talal Aljudi, Samia Esmaeil, Amr Jamal, Alia Habash, Fahad Alsohime, Khalid Alhasan, Ayman Al-Eyadhy

## Abstract

**Background:** Safeguarding children from unintentional injuries is a significant concern for parents and caregivers. With children staying more at home during the COVID-19 pandemic, more educational tools and valid educational programs are warranted to improve parental knowledge and awareness about children’s safety. This study aims to explore the effectiveness of child safety campaigns on parents’ knowledge and attitude towards preventable childhood injuries.

**Methods:** This was a pre-post experimental study, in which the pre-designed assessments were used as an evaluation tool before and after attending a Child Safety Campaign. The Pre – Post assessment question included questions to evaluate the socio-demographic status, followed by knowledge questions in line with the current child safety campaign. The outcomes of interest were assessed before and after attending the campaign’s stations.

**Results:** Three hundred and eight parents volunteered to participate in this study. Their knowledge score improved from 36.2 (SD 17.7) to 79.3 (SD 15.6) after attending the Child Safety Campaign (t-value= 34.6, p<0.001). Both, perceptions on the preventability of accidents and the parents’ perceived usefulness of educational campaigns showed improvements, with (t-value =6.3, p<0.001) and (t-value= 3.097, p<0.001), respectively.

**Conclusion:** The educational child safety campaign for caregivers in Saudi Arabia resulted in a significant increase in the overall knowledge and attitudes towards children’s safety. As children are currently staying at home more, additional educational tools and programs are warranted to promote childhood safe practices among parents and caregivers.

## Introduction

Children are naturally vulnerable and curious; therefore, they tend to explore anything and everything around them, unable to distinguish between harmful and safe objects. This might lead to unintentional injuries that most commonly occur in the home environment because they spend most of their time indoors. Some hazards that might cause injury to children include stairs, sharp objects, and toxic products ^1^. Moreover, many of these unintentional injuries can be prevented by the caregivers responsible for providing a safe, habitable, and healthy environment for children and by teaching them principle methods of self-protection. Thus, caregivers should be encouraged to have good knowledge about the right practices that promote child safety.

Unintentional injuries amongst children are of primary concern all over the world. The consequences can be dire, as the injured child may develop permanent disabilities such as skin burns, amputations, fractures; and in the worst case, injuries may lead to death ^2^. A study conducted in the United States showed that 40% of children’s deaths from ages 1-19 years were related to unintentional injuries ^1^; other studies in Bangladesh, Columbia, Egypt, and Pakistan showed that half of the severe injuries that required an emergency visit resulted in disability ^3^. Similarly, a study conducted in Columbia, Maryland, by Dershewitz et al found that the majority of child mortality between the ages of 1 and 14 are associated with accidental causes that account for approximately 2,800 minor injuries, 97 major injuries, and one death in every 29,000 cases ^4^.

The statistics indicate that the frequency of injuries is high, as unintentional childhood injuries might happen anywhere, like at home, school, in the car, on the road, or in a public area. For instance, a study in Italy showed that most unintentional injuries occurred at home (45.4%), followed by on the road (24.3%), and then at sports facilities (20.3%) ^5^.

However, many injuries, and there consequences, are preventable. To reduce the risk of accidents and injuries, parents and caregivers need to utilize different self-protection measures and provide a safe environment ^6^. Additionally, one of the most effective prevention techniques is through community awareness campaigns. For instance, in the United States, campaigns held to increase awareness about the use of child’s booster seats in motor vehicles showed a significant increase in their use ^7^.

Despite that child safety is a high priority for caregivers, there was no study conducted in Saudi Arabia about the effect of childhood safety campaigns on parents or communities. However, this study aimed to measure and analyze the effect of child safety campaigns on the parents’ knowledge and attitudes.

## Methodology

This was a pre-post experimental study, in which the predesigned assessments were used as an evaluation tool during the Child Safety Campaign (14-17 March 2017). The study assessed its impact on knowledge, attitude, and intended practices among parents in Riyadh, Saudi Arabia.

The targeted population was parents who attended this campaign. The study sample consisted of parents (for children up to 17 years of age and living in Saudi Arabia for at least one year), excluding adults with no children living with them. As no previous data exists locally, a convenience sampling technique was used. We incorporated close-ended questions into an electronic format (using Survey Monkey). The questions were piloted among ten parents to ensure clarity. Then the assessment questionnaire was modified accordingly and tested for validity and consistency before using it in the Child Safety Campaign.

The main components of the questionnaire were sociodemographic data information including: parents’ ages, parents’ education, current employment, current relationship status, and the number of their children. This was followed by knowledge questions about child safety that were in line with the current child safety campaign and parents were tested before and after attending the campaign’s stations.

## Data collection methods

Parents who attended the Child Safety Campaign were invited to participate in this survey. Those who consented to participate in this study were interviewed with the pre-campaign structured questionnaire-A (the bilingual Arabic/English version is available by contacting the corresponding author).

Questionnaire-A consisted of a series of questions that measured their essential demographic and economic characteristics, alongside their perceived self-rating on childhood safety knowledge and their perceptions on the likelihood of preventing childhood injuries.

The parents were also assessed for knowledge on child safety using predesigned questions twice (at baseline and after attending the series of educational materials at the campaign). Parents answered the post-campaign assessment tool (questionnaire-B, the bilingual Arabic/English version is available by contacting the corresponding author), evaluating the same previously assessed aspects, focusing on possible changes in knowledge and attitude towards child safety. The knowledge questions were reshuffled and paraphrased with two additional questions to help alter the questions and avoid recall bias.

Each questionnaire was headed with a letter stating that participation was voluntary, and no identification data were required. Ethical approval was granted by the Institutional Review Board of College of Medicine, King Saud University (Riyadh, Saudi Arabia).

The analysis was performed using Statistical Package for the Social Sciences (SPSS) v19. Basic descriptive analysis was utilized to calculate the frequency and proportion of study variables. Means and standard deviation were calculated to describe continuous variables.

## Results

A total of 308 parents volunteered to participate during the child safety campaign and completed the first set of questions (Questionnaire A) before attending the campaign. Thereafter, the same sample were followed up with linked Questionnaire B.

Out of the participating 308 parents, the majority were mothers (68.8%), and the majority of them were aged above 40 years (79.2%) and married (93.8%). Most of them were educated with a college degree or higher (74%). Saudi nationals comprised the majority of the sample (67.2%) while many of them had monthly income greater than 10,000 SAR per month (36%). The mean number of children for the whole sample was 3.3 children. Reportedly, the primary caregiver at home was the mother by most respondents (93.6%) (Table 1).

**Table 1:** Respondents’ characteristics.

|  | **Frequency** | **Percentage** |
| --- | --- | --- |
| **Sex of the respondent** |  |  |
| Mother | 212 | 68.8 |
| Father | 96 | 31.2 |
| **Age** |  |  |
| Below 40 | 244 | 79.2 |
| Above 40 | 64 | 20.8 |
| **Marital Status** |  |  |
| Married | 274 | 93.8 |
| Widowed | 7 | 2.4 |
| Divorced | 11 | 3.8 |
| **Educational Level** |  |  |
| Elementary/Others | 17 | 5.5 |
| Intermediate | 9 | 2.9 |
| High school | 54 | 17.5 |
| College or higher | 228 | 74 |
| **Nationality** |  |  |
| Saudi | 207 | 67.2 |
| Other nationalities | 101 | 32.8 |
| **Employment** |  |  |
| Employed | 276 | 89.6 |
| Unemployed | 9 | 2.9 |
| Not applicable | 8 | 2.6 |
| Retired | 15 | 4.9 |
| **Income** |  |  |
| No answer | 74 | 24 |
| Below 5000 SAR* | 23 | 7.5 |
| 5000 to 10000 SAR | 100 | 32.5 |
| >10000 SAR | 111 | 36 |
| **Number of children, mean (SD).** | 3.3 (2.2) |  |
| **Principal Child Caregiver** |  |  |
| Mother | 278 | 93.6 |
| Father | 3 | 1 |
| Housemaid | 9 | 2.9 |
| Others | 7 | 2.3 |

\* SAR: Saudi Riyals

The “knowledge questions” used to assess participants’ information on childhood safety are shown in table 2.

**Table 2:** Proportions of correctly answered knowledge questions before and after the educational campaign.

| Knowledge questions | <u>Before Session</u> | <u>After Session.</u> | p |
| --- | --- | --- | --- |
|  | n (%) Correctly answered | n (%) Correctly answered |  |
| Getting rid of old medications is the best way to prevent child medication poisoning | 107 (34.7%) | 220 (71.4%) | <0.001 |
| Hot water at 60 degrees can cause burns; as such heaters should always be set to 60 degrees or below. | 61 (19.8%). | 292 (94.8%) | <0.001 |
| The first life-saving measure for a child with an ingested battery is to take them to the hospital immediately | 130 (42.2%) | 185 (60.1%) | 0.228 |
| IPad devices must remain outside the bedroom of a child when asleep. | 42 (13.6%) | 256 (83.1%) | <0.001 |
| Children May Not be allowed to use electronic devices (e.g., iPad, play stations) for more than 1-2 hours per day. | 205 (66.6%) | 269 (96.1%) | <0.001 |
| Excessive use of social media leads to obesity among children. | 124 (40%) | 256 (83.1%) | <0.001 |
| The appropriate size of the school bag for the weight of the child should not exceed 15% of the child weight | 162 (52.6%) | 257 (83.4%) | <0.001 |
| Your child's school back-bag should be placed on their mid-back when carrying them. | — | 288 (93.5%) | — |
| When using bags with wheels, it is preferred to choose bags with large wheels. | 41 (13.3%) | 190 (61.7%) | <0.001 |
| The appropriate place to install a child seat in the car is the rear-facing back seat. | 159 (51.6%) | 290 (94.2%) | <0.001 |
| A child can use the car's regular seat and seatbelt When the waist belt is in the top level for thighs and the shoulder belt at the chest level. | — | 171 (55.5%) |  |
| The infant in the car must be placed in the back seat in a rear-facing seat when driving | 159 (51.9%) | 209 (67.9%) | <0.001 |
| A fundamental rescue task for the drowning child is doing CPR immediately. | 89 (28.9%) | 267 (86.7%) | <0.001 |

The binomial test showed that a significant proportion of correctly answered questions were observed after the campaign compared to before for most of the used questions with a p-value <0.001, except to whether the first life-saving measure for a child with an ingested battery is to take them to the hospital immediately, p=0.228.

Surveyed parents had significant improvement in their knowledge regarding most of the domains associated with childhood safety post-campaign, whether life-threatening ones as poisoning or drowning, or issues related to IT, and issues related to even skeletal growth and appropriate school bag size handling. For further details refer to Table 2.

For example, their knowledge of getting rid of expired medications to prevent poisoning improved from 34.7% to 71.4%, hot water of 60 degrees Celsius as a cause of burns from 19.8% to 94.8%.

Additionally, the campaign had significantly improved parental knowledge of the correct way of using modern information technology and telecommunication devices that are already integrated into most children’s daily lives. Therefore, the caregiver’s knowledge of the “healthy” way of dealing with electronic devices, their relation to obesity, and the safe amount of screen time, have improved significantly post-campaign.

Participating parents were asked to self-rate their knowledge about childhood safety on a Likert scale of 1-10 before attending the educational campaign; people’s self-rated knowledge was equal to 6.8 points out of 10, which is equal to a scored knowledge (in marks out of a maximum 100) of 36.2 points at the baseline. However, their knowledge score recorded a sharp rise after the campaign educational sessions were delivered; their overall knowledge score was 79.3 points out of 100 with a p-value <0.001.

Their belief in preventing unintentional child injuries and their perceived usefulness of childhood safety campaigns increased significantly post-campaign, as shown in table 3.

**Table 3:** Parental belief in childhood injuries preventability and perceived usefulness of childhood safety campaigns.

|  | Before session | After Session |  |
| --- | --- | --- | --- |
|  | mean(SD) | mean(SD) | p |
| Belief in Childhood safety prevention (%) | 67.6 (22.9) | 76.3 (20.1) | <0.001 |
| Perceived usefulness of Childhood safety Campaigns (1-10 score). | 8.8 (1.7) | 9.1 (1.2) | <0.001 |
| Knowledge score (%). | 36.2 (17.7) | 79.3 (15.6) | <0.001 |

## Discussion

The number of deaths among children younger than five years has declined substantially over the past 47 years, and this is evidence that progress is being made in tackling the fundamental causes of childhood mortality^8^. An example of a successful strategy includes increasing the educational levels of mothers^9^. Still, unintentional injuries are a worldwide health problem that causes high death rates among children younger than 18. Furthermore, they are the most common cause of hospital admissions and permanent disabilities ^10^.

The spectrum of unintentional injuries is diverse, including ingestion of toxic materials such as medications or poisoning materials, foreign bodies ingestion or aspiration, falls, traffic accidents, or burns ^11–13^. Many of these injuries could be prevented if parents and caregivers have good literacy about different types of injury and preventative measures and understand the health information and instructions to deal with emergency matters that may result ^14 15^. However, caregivers may not be adherent to safety practices and the recommended injury prevention tasks. Therefore, continued emphasis on these strategies is recommended ^16 17^.

The present study showed that before the implementation of the educational program, most caregivers had insufficient knowledge about the preventative measures of childhood safety issues, including the seatbelt, discarding expired medications and batteries, using very hot water, the appropriate way to use electronic devices, using an appropriate car seat, and first aid for a drowning child. International papers have reported similar unsatisfactory outcomes, emphasizing the need for global and nationwide interventional educational campaigns to correct the current knowledge deficit ^18 19^.

No previous studies in Saudi Arabia evaluated the knowledge and attitude among caregivers regarding child safety or assessed the impact of educational safety campaigns on the caregivers’ overall knowledge and practices. After the intervention of child safety campaigns, the overall knowledge and perception of the usefulness of campaigns and preventability of injuries were significantly increased. Thus, educational campaigns about child safety measures could result in increasing the KAP of caregivers, and in decreasing the rate of injuries, permanent disabilities as well as child death. A study conducted in Brazil observed an increase in mothers’ level of knowledge after the educational intervention was applied compared to their basic prior knowledge, confirming the importance of implementing community awareness programs ^1^.

Educational interventions have shown their tremendous benefits in promoting the level of knowledge and awareness among caregivers and limiting unintentional childhood injuries. Therefore, we strongly acknowledge the need for combined efforts of all concerned institutions to establish a national awareness program that can reinforce best practices, reduce ineffective measures and ensure children’s safety. Many countries have inserted these educational campaigns as a powerful program to increase the protection of children and reduce mortality rates ^10 20 21^.

Several nations have decreased the rates of injury-related deaths in more than 50% of children through educational safety programs ^22^. As such, health authorities should establish injury prevention programs like road accidents prevention and clear child restraint laws. One of the attempts in this regard was the Brazilian National Policy for Reduction of Mortality from Accidents and Violence established by the Brazilian Ministry of Health ^1^. Other educational training programs for children’s first aid are recommended for all subjects to rescue drowning children. While many Saudi cities are located in a desert-environment, drowning could still occur in recreational swimming pools ^23^. The Saudi Red Crescent Authority has a program called Prince Naif first aid program, which targets all population groups, and could be utilized to improve parents’ first-aid skills ^24^.

Other strategies to alleviate harm from burns were successful, such as using smoke alarms, safer lamps, and laws on the temperature of hot-water taps, resulting in decreased incidence of disabilities and deaths from burns during childhood ^25^.

The death count from poisoning by chemical and medical substances could be prevented by sufficient parental supervision and safe storage of such substances while discarding hazardous ones ^15 26^. With the COVID-19 pandemic, some agencies, like the Consumer Product Safety Commission in the USA, published Home Safe Checklists, so parents and caregivers could check off the safety items in their home environments ^27^.

While our study was the first to explore the impact of a child safety campaign amongst parents in Saudi Arabia, it still had some limitations. The self-reported practices may not reflect actual behaviors but remain among the best available tools to assess the population’s knowledge, attitude, and reported practices. The use of the same reporting tool twice (pre-post campaign changes) may have added carried-on bias to some participants, though the questions were rephrased to minimize this possibility. Future longitudinal studies that follow the participants are warranted to seek whether these changes are maintained over time; and whether these improvements in parental knowledge and attitudes are translated into better child safety practices.

## Conclusion

The educational child safety campaign for caregivers in this study resulted in a significant increase in the overall knowledge and awareness of children’s safety and preventable measures. As children stay home more during the COVID-19 pandemic, more educational tools and programs are warranted to promote child safety practices among parents and caregivers.

## Data Availability

All the data for this study will be made available upon reasonable request by emailing

## Declaration

### Ethics approval, guidelines and consent to participate

Approval from the Institutional Review Board at King Saud University, Riyadh, Saudi Arabia, was obtained before conducting the study. We confirm that all methods were performed in accordance with the relevant guidelines and regulations. The informed consent was obtained from all subjects prior to enrollment. No identifiable data was collected, and participants were assigned code numbers to maintain confidentiality.

### Consent for publication

All authors gave their consent for publication.

### Availability of data and materials

All the data for this study will be made available upon reasonable request by emailing to

### Authors’ contributions

MHT, FAJ, AAE, BA, and RA conceptualized the study, analyzed the data, and wrote the manuscript. FAS, AAH, KAH, TA, AF, AS, and RT contributed to the study design; collected, analyzed, and interpreted data; and edited the manuscript. MHT contributed to the study design and interpretation and edited the manuscript. AJ, SE, AH interpreted the data and finalized the manuscript. All authors reviewed and approved the final version of the manuscript.

## Acknowledgements

This research has been financially supported by Prince Abdullah Ben Khalid Celiac Disease Research Chair, under the Vice Deanship of Research Chairs, King Saud University, Riyadh, Kingdom of Saudi Arabia. We would like to thank hodhodata.com for their statistical analysis.

